# Malnutrition and risk of kala-azar: a systematic review with meta-analysis

**DOI:** 10.1101/2025.07.10.25330979

**Authors:** Daniele Vieira da Silva Blamires, Dorcas Lamounier Costa, Carlos Henrique Nery Costa

## Abstract

This systematic review was conducted with the objective of clarifying the association between malnutrition and the risk of developing kala-azar in humans. The research question was formulated using the PECO acronym: “Is malnutrition a risk factor for the development of kala-azar?” The search was carried out between September and November 2022, with an update in October 2024, in the PubMed, Embase, CAPES Journal Portal databases, and the grey literature. Original studies including individuals exposed to kala-azar with an evaluation of their nutritional status were included. Of the 1,261 records identified, five studies met the inclusion criteria, comprising four cohort studies and one case-control study. Four studies evaluated the association between malnutrition and clinical disease, and one evaluated the association with asymptomatic infection. Only two studies demonstrated a statistically significant association between malnutrition and kala-azar. A meta-analysis was conducted with two studies using R Studio software. The relative risk found was higher for malnourished individuals, although without statistical significance, possibly due to the high heterogeneity among the studies. A higher risk of kala-azar was observed in malnourished children, although without statistical concordance among findings. The methodological quality of the studies was considered low, with a high risk of bias, especially regarding the classification between primary and secondary malnutrition. It is concluded that there is evidence suggesting an association between malnutrition and kala-azar, but it is not possible to affirm this causal relationship with certainty. Further studies with greater methodological rigor, longer follow-up, and larger sample sizes are needed to confirm whether malnutrition modifies the risk of developing kala-azar.

## INTRODUCTION

The definition of malnutrition adopted by the World Health Organization encompasses deficiencies, excesses or imbalances in energy or nutrient intake, covering three major groups: malnutrition, micronutrient deficiency and overnutrition. Malnutrition can be classified as acute (wasting), chronic (stunting), or a combination of both (underweight). Micronutrient deficiency refers to the insufficiency of vitamins and minerals. The classification of malnutrition also includes overweight, obesity, and diet-related noncommunicable diseases such as heart disease, stroke, and some cancers ^1^.

Wasting malnutrition is defined by low weight in relation to height and can also be measured by arm circumference. Stunting is shortness in height relative to age, and the combination of the two, underweight, is low in weight relative to age. Malnutrition can also be classified as mild, moderate or severe, according to the nutritional reduction in relation to a reference population, measured by the number of standard deviations (*z-* score) in relation to the measurement of the central tendency; when the combinations of anthropometric measures of weight, height and age are one or two standard deviations below the reference median, mild or moderate malnutrition is considered ^2–4^. Brachial circumference greater than 115 mm or less than 125 mm indicates moderate acute malnutrition. Severe malnutrition, with high lethality, includes marasmus, caused by lack of energy, with a *z*-score below three standard deviations from the median, or brachial circumference less than 115 mm, and kwashiorkor, characterized by lack of proteins, leading to fluid retention, edema and skin changes ^5^.

In addition, malnutrition can be primary, due to lack of energy intake or absorption, associated with factors such as poverty, conflicts or natural disasters, or secondary, caused by pathophysiological disorders from underlying diseases that affect growth and compromise appetite, nutrient absorption and catabolism ^6^. In 2022, 148.1 million children under the age of five had stunting (22.3%) and 45 million suffered from wasting (6.8%). These numbers underestimate the reality, since many children with nutritional edema due to kwashiorkor were not accounted for ^7^. Malnutrition weakens the immune response, increasing the risk of infectious diseases, by various mechanisms such as increased enteric permeability, reduced humoral and cellular immunity, and decreased thymus size ^8–10^.

This study examines the causal relationship between malnutrition and kala-azar, or visceral leishmaniasis, a parasitic disease that affected between 50,000 and 90,000 people in 2022. If left untreated, most patients progress to death ^11^. Kala-azar is an anthroponosis caused by protozoa of the genus *Leishmania*, being *L. donovani* the main cause in South Asia and East Africa, while *L. infantum* is predominant in the Americas, Central Asia and the Mediterranean basin, where it is a zoonosis transmitted by sandflies, with high mortality among dogs and significant morbidity among humans, especially children and adults infected by the HIV or other immunodeficiencies ^12^.

Research indicates that a significant part of the population in non-endemic areas has previous *Leishmania* infection, without clinical symptoms, suggesting that the vast majority of infections do not progress to kala-azar ^13–18^. In humans, early symptoms include inappetence, pallor, weight loss, increased abdominal volume, and low-grade fever, followed by malnutrition, anemia, and prolonged fever, which lead patients to seek medical attention. The diagnosis is confirmed by laboratory tests ^19^. Even with treatment, about 10% of patients do not survive, with a high mortality rate in Brazil ^20,21^.

*Leishmania* infection results in a gradual fight between the parasites and the immune system. Cellular immunity is essential to control the disease, and in patients with AIDS or other immunosuppression, the infection becomes more aggressive ^22^. Since malnutrition weakens immune defenses, malnourished people in endemic areas are believed to be more vulnerable to the disease. This study seeks to synthesize the current knowledge about the relationship between malnutrition and kala-azar, with a systematic review and meta-analysis of the studies conducted.

The systematic review was conducted following the guidelines of the Cochrane Collaboration and the Ministry of Health ^23^. A protocol was registered in PROSPERO (ID CRD42023413416) and aimed to answer the research question: “Is malnutrition a risk factor for the development of kala-azar?” using the acronym PECO (Population: Population exposed of kala-azar, Exposure: Malnutrition, Comparator: Eutrophic people and people with malnutrition, Outcomes: Diagnosis of kala-azar or Asymptomatic infection).

## METHODS

Original studies with individuals exposed to kala-azar and with assessment of nutritional status were included. There were no restrictions on the period of publication, language, gender, age or socioeconomic status, but studies with participants already diagnosed with kala-azar during nutritional assessment were excluded.

Study identification data were collected, such as title, year of publication, study design, follow-up period, and place where the study was conducted. The characteristics of the participants, methods of assessing malnutrition and kala-azar, and results obtained were also recorded. Narrative synthesis included the methodological characteristics of the studies and the exposure and effect measurements.

The bibliographic search was carried out in PubMed, EMBASE, CAPES Journals Portal, as well as theses and dissertations catalogs from CAPES and the Brazilian Digital Library. The research took place in October 2024. Controlled vocabularies such as DeCS, MeSH, and Emtree were used to identify relevant descriptors. The search strategy was: (((visceral leishmaniasis) OR (kala azar)) AND ((nutrition) OR (malnutrition)) NOT ((HIV) NOT (AIDS) NOT (immunodeficiency))).

The results were exported to the Rayyan tool (https://www.rayyan.ai/), for duplicate removal. The selection of studies followed two phases: in the first, studies were screened based on titles and abstracts; in the second, the reviewers obtained the complete reports for detailed analysis. Two reviewers (DB and CH) performed the selection and extraction of data independently, with resolution of disagreements by consensus or by consultation with a third reviewer (DL).

A meta-analysis was performed with two studies, using the R Studio software (version 2023.09.1). In one study, the effect of moderate to severe underweight was compared with the nutritional status of eutrophic or mildly malnourished patients. In the other, the effect of moderate to severe stunting was compared with that of eutrophic patients.

Each study was assessed for risk of bias using the Research Triangle Institute Bank (RTI) critical assessment list, supplementary table S1,S2. To adjust the number of words to that required by the journal, we prompted ChatGPT to “Reduce to x words keeping the words in parentheses” in the introduction, material and methods and results, followed by manual correction. In the last five paragraphs of discussion, we prompted the AI to “Reduce by keeping words in parentheses”, followed by manual correction.

## RESULTS

1.261 references were identified through the search strategies described in the methods. After removing duplicates, 989 publications were selected for screening by title and abstract. At the end of this stage, 10 titles were chosen for full-text evaluation. After the last eligibility analysis, five studies were included in the review: four cohort studies and one case-control study. The process of identification, screening, and eligibility assessment is summarized in Figure 1.

**Figure 1.**
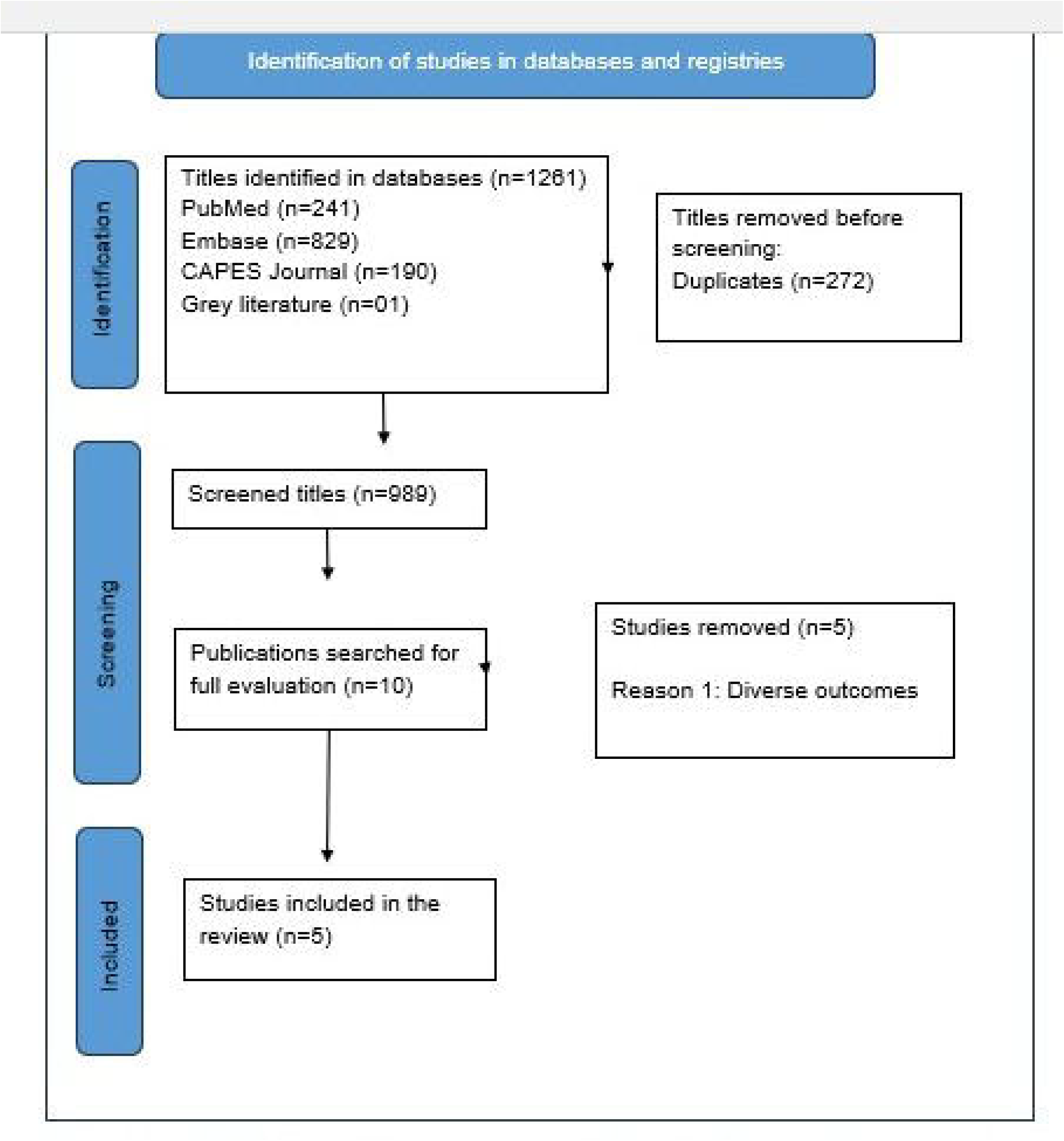
Flowchart of the eligibility process and identification of studies of publications tracked in databases and records

Table 1 presents the overall characteristics of the studies, including the distribution of specific characteristics, such as the measurement of exposure and effect, the time of disease evolution and the association between kala-azar and nutritional status. The sample ranged from 136 to 1,320 individuals, all living in the Northeast region of Brazil (States of Ceará, Bahia and Maranhão). In all studies, the exposed population was composed mainly of children. Cohort studies included ages as follows: up to 5 years old; from 10-12 months to 11 years old; and up to 15 years old. The case control study included control participants up to 44 years old. The diagnosis of kala-azar was performed by serology and parasitological examination of bone marrow aspirate, while intradermal tests and serology were used to identify asymptomatic infection.

**Table 1.** Heterogeneity of the studies: study design, expo sine measurement, comparison and classification of the degrees of exposure, and evaluated outcome.

| Authors | Type of Study | Exposure measurement | Comparison of degrees of exposure | Classification of degrees of malnutrition | Types of outcomes |
| --- | --- | --- | --- | --- | --- |
| Harrison et al., 1986 | Case-control study | Skin fold<br>Arm circumference | Measurement averages | Not done | Disease |
| Cerf et al., 1987 | Cohort | Height-for-age (stunting)<br>Weight-for-age (underweight) | Proportions: eutrophic/mild vs moderate/severe | Relative proportion (%) to reference | Disease |
| Evans et al., 1992 | Cohort | Weight-for-height (wasting)<br>Height-for-age (stunting)<br>Weight-for-age (underweight) | Average standard scores | Z scores | Disease |
| Nascimento, 1996 | Cohort | Weight-for-height (wasting)<br>Weight-for-age* | Proportions: eutrophic/mild vs moderate/severe | Z scores to reference | Disease |
| Caldas et al., 2002 | Cohort | Height-for-age (stunting) | Proportions: eutrophic/mild vs moderate/severe | Z scores to reference | Asymptomatic infection |

\* Uncertain classification. \*\* Mild malnutrition was considered eutrophic.

Three measures of malnutrition were analyzed. Two were based on reference values of the United States population and an assessment of nutritional status was made with anthropometric measurements of the arm (triceps skinfold and brachial circumference), in addition to weight, height and age. Children with weight-for-age between 75% and 90% of the reference median were classified as mildly malnourished, while those with less than 75% of weight-for-height or height-for-age were classified as moderately to severely malnourished. Another assessment used the *z*-score and its standard deviation to define moderate to severe malnutrition.

The studies were published over the course of 16 years, with the oldest having been published 23 years ago, which makes them relatively outdated ^24^. All studies were longitudinal, with possible causal inferences. One study was case-control and the other four were cohort studies, one of them being an open cohort. The assessment of nutritional status varied between studies, which made a direct comparison difficult. The oldest case-control study ^24^ used brachial circumference and triceps skinfold to classify children, while the cohort studies used weight, height, and age. In all studies, malnutrition was associated with the incidence of kala-azar or asymptomatic infections.

The study by Harrison *et al*. ^24^ found that cohabitants of the same household as children with kala-azar had 22% less body fat compared to neighborhood controls. The study by Cerf *et al*. ^25^ showed that 77% of children with kala-azar had acute or chronic malnutrition before symptoms, compared to 53% of uninfected children (*p* < 0,05). This study also revealed that children with moderate to severe malnutrition had an 8.7x higher risk of developing kala-azar than eutrophic children. However, this study did not provide data on the time of onset of disease symptoms.

The study by Evans *et al*. ^17^ verified moderate to severe malnutrition in 32% of the children, but did not verify statistical significance in the results. This study, which investigated early symptoms of kala-azar, was the most rigorous in relation to early monitoring of signs of the disease, but did not allow to calculate the association between malnutrition and the risk of developing disease due to the lack of data on the proportions of exposed children who developed kala-azar.

In Nascimento’s ^26^ study, no significant association was observed between malnutrition and kala-azar, despite a difference in risk between children with acute and eutrophic malnutrition. The study by Caldas *et al*. ^27^ investigated chronic malnutrition and asymptomatic infection; although malnutrition was associated with a higher incidence of infection, the results were not declared statistically significant.

Table 2 summarizes the associations between types of malnutrition and outcomes. The study by Harrison *et al*. ^24^ revealed a significant increased risk of kala-azar in malnourished children. The studies by Evans *et al*. ^17^ and Nascimento ^26^ found no statistically significant associations between acute malnutrition and kala-azar. The study by Cerf *et al*. ^25^ found a significantly higher risk of kala-azar in children with chronic malnutrition. Cohort studies looked at the combination of acute and chronic malnutrition, identifying an increase in risk but with inconsistent results.

**Table 2.** Risk of asymptomatic infection with Leishmania sp. according to the previous nutritional assessment, per study analyzed.

| Study | Unspecified malnutrition | Acute malnutrition | Chronic malnutrition | Acute and chronic malnutrition | Acute or chronic malnutrition |
| --- | --- | --- | --- | --- | --- |
| Harrison et al., 1986 | Increased risk<br>$p < 0.05$ | Not evaluated | Not evaluated | Not evaluated | Not evaluated |
| Cerf et al., 1987 | Not evaluated | Not evaluated | Increased risk<br>$p = 0.02$ | Increased risk<br>$p < 0.05$ | Not evaluated |
| Evans et al., 1992 | Not evaluated | Increased risk, but not significantly | Increased risk, but not significantly | Increased risk, but not significantly | Not evaluated |
| Nascimento, 1996 | Not evaluated | Increased risk, but not significantly | Not evaluated | Not evaluated | Increased risk, but not significantly |
| Caldas et al., 2002 | Not evaluated | Not evaluated | Increased risk, but not significantly<sup>1</sup> | Not evaluated | Not evaluated |

<sup>1</sup>After redoing the statistical calculations, significance was found ( $p < 0.05$ ).
<sup>2</sup>The authors did not clarify whether they refer to low weight-for-age only or to combined indicators.

A meta-analysis was performed with two studies that presented similar methodological characteristics, as detailed in Figure 2. The pooled analysis of the studies indicated that moderate to severe malnutrition was not associated with an increased risk of kala-azar compared to eutrophic children, with a relative risk of 3.7 (95% confidence interval: 0.56; 24.35). The heterogeneity between the studies was high (I^2^ = 82%, *p* = 0.02), suggesting that the studies were excessively heterogeneous to be evaluated together.

**Figure 2.**
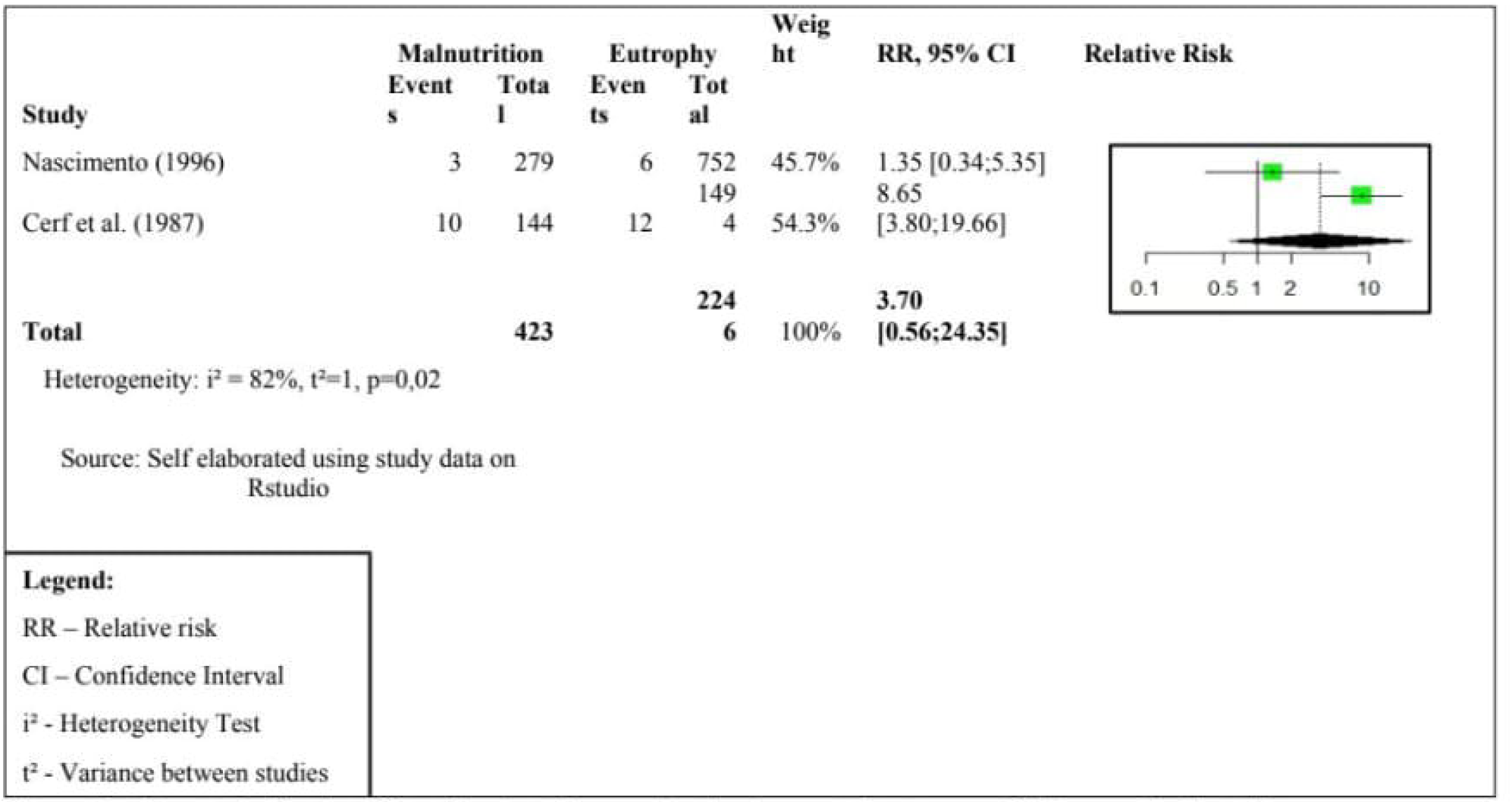
*Forest plot* for the effect of malnutrition on the developmental outcome of kala-azar. Both studies evaluated the effect of malnutrition on the incidence of the disease.

The risks of bias were considered high when assessed using the RTI tools, due to the lack of investigation of confounding factors, lack of strategies to deal with incomplete follow-up, and lack of information on follow-up time. The certainty of the evidence was considered very low, mainly due to methodological limitations, inconsistency (high heterogeneity) and imprecision (wide range of the confidence interval).

## DISCUSSION

Overall, individual studies have shown a trend of association between malnutrition and increased risk of asymptomatic infection by *L. infantum* progressing to the development of kala-azar, although only two studies have recorded statistically significant increases ^24,25^. This discrepancy suggests that at least one of three possible phenomena occurred in the studies analyzed: (a) the effect actually exists, but the size of the samples was not sufficient to distinguish the effects of malnutrition on the incidence of kala-azar (type II error); (b) the effect does not exist because there was bias indicating associations that, in reality, did not exist (type I error); (c) a confounding variable was present that was not controlled.

Regarding statistical power, it is possible that there was a type II error in the study by Evans *et al*.^17^, as the authors themselves admitted, and also in the study by Nascimento ^26^. In the latter, the data in Table 2 of this study suggest that the statistical power may have been insufficient to distinguish the slight differences in incidence observed.

An exposure classification bias may have been caused by the ambivalent cause-and-effect association between malnutrition and kala-azar. The problem was aggravated by the fact that the first symptoms of kala-azar, such as inappetence, weight loss and pallor, are often barely noticeable and can last a long time before the disease is noticed. In this situation, by the temporal order of events, malnutrition could be mistakenly interpreted as primary ^28^ when in fact it could have been secondary to the disease itself, falsely suggesting that malnutrition is a risk factor for kala-azar. In addition, the prodromal period of kala-azar is not well defined. In fact, no results were found in a PubMed search conducted in August 2024, using the search strategy ((visceral leishmaniasis) OR (kala-azar)) AND (prodrome)).

In addition, the description of cases of subclinical infection suggests that the prodromal period of the disease is considerably long ^16^. In addition, it is plausible that patients with a longer prodromal period present more pronounced weight loss due to anorexia and increased lipolysis and protein catabolism, resulting from the action of

pro-inflammatory cytokines, such as TNF-α ^29^. This argument is reinforced by the fact that malnutrition secondary to disease can last for months after cure, as observed in the studies by Harrison *et al* and Evans *et al* ^17,24^ Active investigation of constitutional symptoms could reduce this bias, as was done in the study by Evans *et al*.^17^, albeit qualitatively and potentially inaccurately. The study by Harrison *et al*.^24^, however, did not allow the control of this bias.

Finally, age may have been a confounding variable, as it is associated with both malnutrition and the incidence of kala-azar; as both associations are negative, the tendency would be to overestimate the risk. In the study by Nascimento ^26^ (1996), although a multivariate analysis was performed to control the effect of age, the adjusted risk was not shown in the publication.

The study by Cerf *et al*. ^25^ showed a greater potential misclassification bias of exposure, due to the longer period between nutritional assessment and diagnosis of kala-azar, while the study by Evans *et al* (1992) showed a greater risk of type II error. The combination of acute and chronic malnutrition also presented conflicting results between the studies by Cerf *et al*. ^25^ and Evans *et al*. ^17.^ However, a third study found no significant association between this more severe type of malnutrition and the evolution of infection by *L. infantum* to kala-azar ^26^.

Only four studies were eligible for the meta-analysis. One of them, however, had an asymptomatic infection as an outcome and another did not compare the incidence of the disease. Thus, two cohort studies whose outcome was the incidence of kala-azar were included. Although both indicated higher risk, the results were quite discrepant. One of the reasons for this was the difference in the evaluated exposures: while the study by Cerf *et al*. ^25^ measured the effect of underweight (low weight-for-age), the Nascimento study^26^ evaluated wasting (low weight-for-height). As the underweight effect was much more pronounced, it is plausible to conjecture that underweight affects host defenses more than wasting because, while wasting refers to acute malnutrition, reflecting relatively recent events, underweight represents a chronic malnutrition that has recently been aggravated, which may more seriously compromise the cells and tissues of the immune system.

The case of the publication by Caldas *et al*. ^27^ is peculiar in relation to the occurrence of type II error. The study concluded that there was no statistically significant difference between eutrophic and chronically malnourished children regarding the probability of asymptomatic infection, measured by both cellular immunity and humoral immunity. However, when redoing the statistical analysis in this review, a different result was found.^a^ Table 3 of the work by Caldas *et al*. ^27^ shows that, over seven months of observation, children with chronic malnutrition had an incidence of positivity in the skin test of 14/132 (10.6%), compared to 13/337 (3.9%) among eutrophic children, generating a relative risk of 2.7, with *p* = 0,01. Regarding serology, the incidence was 31/138 (22.5%) among chronically malnourished children, compared to 34/347 (9.8%) among eutrophic children, resulting in a relative risk of 2.3, with *p* =

0.002. These data suggest that chronic malnutrition is associated with a higher risk of infection by *L. infantum* in endemic areas in Brazil. Two hypotheses may explain this phenomenon. The first is that malnourished people may be more attractive to the *Lutzomyia longipalpis* vector due to the emission of different kairomones in relation to eutrophic people. The second is that chronically malnourished children live in more precarious environments, subject to hunger and prone to the transmission of kala-azar and other infections, as observed in the study by Harrison *et al*.^24^ and in other epidemiological studies ^30^.

Systematic reviews ^9,10^ revealed a set of immunological changes present in moderate malnutrition, such as the reduction of thymus size, the increase of IL-4 and IL-10, and the reduction of IL-2, IL-12 and INF-g, which facilitate the development of opportunistic infections. Even more pronounced changes were described in severe malnutrition. Unfortunately, the discrimination between acute and chronic malnutrition was not made in the aforementioned reviews, making it difficult to formulate hypotheses about the immunosuppressive mechanisms associated with malnutrition.

We also sought to investigate whether distinct types of malnutrition could have distinct effects on the occurrence of opportunistic infections. Acute malnutrition indicates recent weight loss, resulting from either reduced intake or absorption, or consumptive diseases. This is a serious problem in which young children are at increased risk of mortality ^31–33^. The occurrence of severe drought followed by widespread hunger in Northeast Brazil preceded and seems to have triggered the first studies analyzed in this systematic review ^34^ before supplemental food programs were created in Brazil. An autopsy study of people who died from hunger strike describes the devastating and widespread effect of primary acute malnutrition ^35^. Unfortunately, no comparative studies are available between the effects on immunity of acute and chronic malnutrition. The latter is the result of prolonged or repetitive malnutrition processes, associated with precarious socioeconomic conditions, inadequate maternal health and nutrition, frequent diseases and insufficient care in the feeding of young children, delaying physical development and impairing cognitive potential^3^. As low weight-for-age represents a combination of acute and chronic malnutrition, it is supposedly more severe and threatening than acute or chronic malnutrition alone, as it acutely affects already chronically malnourished individuals, potentially leading to greater impacts on immunity and facilitating the occurrence of opportunistic infections. The data from the studies in this systematic review supports this suggestion.

Some studies indicate that the proportion of infected people who progress to the clinical manifestation of the disease is less than 1% ^30^. However, this proportion seems to be significantly higher among people with some type of immunodeficiency ^22^, which allows the classification of kala-azar as an opportunistic disease, as it would be in malnutrition.

In this initial interactive environment, peptides presented to CD4+ and CD8+ lymphocytes via major histocompatibility complex molecules can trigger different responses. These responses can determine a cellular adaptive response, with Th1 type CD4 lymphocytes inducing activation of infected macrophages through secretion of INF-g, TNF-a, and IL-12, resulting in infection containment and generally, permanent cellular immunity ^36,37^. However, a minority of infected people develop a Th2 type T helper or T-regulator, with suppression of INF-g and predominance of immunoregulatory cytokines, such as IL-4, IL-10 and IL-13. Under these circumstances, phagocytes allow the multiplication of parasites, which migrate to secondary lymphoid organs and tissues, giving rise to the symptoms of LV ^38^.

Previous studies with hamsters have shown that acute protein malnutrition can aggravate infection ^39^. However, the infection model used was acute malnutrition, it was not natural, limiting the interpretation of the results and their generalization to humans. In this systematic review, the two studies that analyzed acute malnutrition ^17,26^ found no significant association with kala-azar. Regarding chronic malnutrition, there was a divergence between the results, as only one of the two studies showed a significant association ^17,25^.

The molecular details that alter the balance of the host response, favoring an immune profile of susceptibility to kala-azar in malnutrition, are still unclear. Thymic atrophy combined with humoral and cellular dysfunctions, such as complement deficiency and effector T cells, may explain the higher incidence of kala-azar in chronic malnutrition ^8–10^. However, the heterogeneity between the studies analyzed limited the conclusions about the effects of acute and chronic malnutrition on kala-azar.

## CONCLUSION

Although malnutrition appears to facilitate the development of kala-azar, this review did not clearly define whether it poses a risk. The question remains open and is of great relevance, requiring further studies to identify the immunobiological phenomena of malnutrition that contribute to the evolution of an asymptomatic infection into the disease. These findings may clarify other conditions related to failure of cellular immunity.

It is important to mention the strengths and limitations of this review. Among the strengths, we highlight the scope of the search and the presence of studies that allowed the combined analysis of robust and accurate association measures (relative risk with confidence interval), favoring the verification of associations between exposure and outcome. However, among the main limitations, the heterogeneity between the studies stands out due to the methodological diversity and the scarcity of studies with strict monitoring in which the participants were not previously infected.

Since the period of the studies reviewed here, the disease has ceased to be restricted to endemic rural areas, HIV co-infection has expanded and new immunosuppressive drugs have emerged, resulting in a new epidemiology of the disease. Simultaneously, food supplementation programs were implemented, reducing the impact of hunger and its effects on kala-azar. Perhaps as a consequence of these changes, there is a decline in scientific interest in the relationship between malnutrition and kala-azar, which is worrying.

Since kala-azar is infrequent after infection, it makes cohort studies with large samples and long duration essential to clarify nutritional and other risk factors, with strict control of symptoms before diagnosis. However, these studies require significant investments, unlikely in the current scenario. However, global warming already affects food production ^40,41^, and the threat of nuclear conflicts may result in global hunger crises ^42,43^. Therefore, understanding the impact of malnutrition on disease occurrence remains a crucial and urgent issue, requiring research on how to mitigate the effects of hunger on infectious diseases.

## Data Availability

All data produced in the present study are contained within the manuscript.

## Footnotes

a The corresponding author and the journal have been informed of the issue.

