## Supplementary material for "Malnutrition and risk of kala-azar: a systematic review with meta-analysis": table suplemetare 1

Supplementary table S1. Assessment of the risk of bias of the studies included by the RTI tool for the cohort studies of Cerf et al., (1987), Evans et al., (1992), Nascimento (1996), and Caldas et al., (2002).

| **List of RTI* critical assessment items – cohort studies** | **Evaluation** | | | |
| --- | --- | --- | --- | --- |
|  | **Cerf et al., (1987)** | **Evans et al., (1992)** | **Nascimento (1996)** | **Caldas et al., (2002)** |
| 1. Is the study design prospective, retrospective, or mixed? | Retrospective | Prospective | Retrospective | Prospective |
| 2. Are inclusion/exclusion criteria clearly stated? | Yes | Yes | Yes | Yes |
| 3. Are inclusion/exclusion criteria validly and reliably measured? | Yes | Yes | Yes | Yes |
| 4. Did the study apply the inclusion/exclusion criteria uniformly? | Yes | Yes | Yes | Yes |
| 5. Was the recruitment strategy the same for all groups? | Yes | Yes | Yes | Yes |
| 6. Was the sample size sufficient? | No | No | No | No |
| 7. Was the exposure/intervention well described? | Average | Average | Average | Average |
| 8. Were the results pre-specified by the researchers? | Yes | Yes | Yes | Yes |
| 9. Was the comparison group well selected? | Yes | Yes | Yes | Yes |
| 10. Was there an attempt to balance the groups? | No | No | No | No |
| 11. Did the study isolate the impact of other exposures? | Yes | Yes | Yes | Yes |
| 12. Did the study follow the planned protocol? | Yes | Yes | Yes | Yes |

| **List of RTI* critical assessment items – cohort studies** | **Evaluation** |  |  |  |
| --- | --- | --- | --- | --- |
|  | **Cerf et al., (1987)** | **Evans et al., (1992)** | **Nascimento (1996)** | **Caldas et al., (2002)** |
| 13. Were the evaluators blinded to the exposure/intervention? | Not applicable | Not applicable | Not applicable | Yes |
| 14. Have interventions/exposures been validly and reliably measured? | Yes | Yes | Yes | Yes |
| 15. Were outcomes assessed with valid and reliable measures? | Yes | Yes | Yes | Yes |
| 16. Was the duration of follow-up the same for all groups? | Yes | Yes | Yes | Yes |
| 17. Was the follow-up period sufficient to evaluate the results? | Yes | Yes | Yes | Yes |
| 18. Did the study have high loss to follow-up (>20-30%)? | Not applicable | Not applicable | Not applicable | Not applicable |
| 19. Was the loss to follow-up different between groups (>20%)? | No | No | No | Yes |
| 20. Did the analysis control for baseline differences between groups? | Not applicable | Not applicable | Not applicable | Not applicable |
| 21. Have confounding variables been validly and reliably measured? | Not applicable | Not applicable | Not applicable | Not applicable |
| 22. Were the confounding variables adjusted correctly in the analysis? | No | No | No | No |
| 23. Has a loss-to-follow impact analysis been done? | Not applicable | Not applicable | Not applicable | Not applicable |
| 24. Were any important primary outcomes missing? | Yes | Yes | Yes | Yes |

| **List of RTI* critical assessment items – cohort studies** | **Evaluation** |  |  |  |
| --- | --- | --- | --- | --- |
|  | **Cerf et al.,**  **(1987)** | **Evans et al.,**  **(1992)** | **Nascimento**  **(1996)** | **Caldas et al., (2002)** |
| 25. Were the statistical methods adequate? | Yes | Yes | Yes | Yes |
| 26. Were any expected adverse events missing from the results? | Yes | Yes | Yes | Yes |
| 27. Was the statistical analysis of adverse events adequate? | No | No | No | No |
| 28. Are the results reliable considering the limitations of the study? | Yes | Yes | Yes | Yes |
| 29. Has the funding source been identified? | No | No | No | No |
| **Risk of general bias** | **High** | **High** | **High** | **High** |
