## Supplementary material for "Malnutrition and risk of kala-azar: a systematic review with meta-analysis": table suplementare 2

Supplementary table S2. Assessment of the risk of bias of the studies included by the RTI tool for the case-control of Harrison et al., 1986.

| **List of critical evaluation items RTI* – case control study** | **Evaluation** |
| --- | --- |
| 1. Is the study design prospective, retrospective, or mixed? | Case control / mixed |
| 2. Are inclusion/exclusion criteria clearly stated? | Yes |
| 3. Are inclusion/exclusion criteria validly and reliably measured? | Yes |
| 4. Did the study apply the inclusion/exclusion criteria uniformly? | Yes |
| 5. Was the recruitment strategy the same for all groups? | Yes |
| 6. Was the sample size sufficient? | No |
| 7. Was the exposure/intervention well described? | Average |
| 8. Were the results pre-specified by the researchers? | Yes |
| 9. **.** Was the comparison group well selected? | Yes |
| 10. Was there an attempt to balance the groups? | No |
| 11. Did the study isolate the impact of other exposures? | No |
| 12. Did the study follow the planned protocol? | Yes |
| 13. Were the evaluators blinded to the exposure/intervention? | Not applicable |
| 14. Have interventions/exposures been validly and reliably measured? | Yes |
| 15. Were outcomes assessed with valid and reliable measures? | Yes |
| 16. Was the duration of follow-up the same for all groups? | Yes |
| 17. Was the follow-up period sufficient to evaluate the results? | Yes |
| 18. Did the study have high loss to follow-up (>20-30%)? | Not applicable |
| 19. Was the loss to follow-up different between groups (>20%)? | No |
| 20. Did the analysis control for baseline differences between groups? | Not applicable |
| 21. Have confounding variables been validly and reliably measured? | Not applicable |
| 22. Were the confounding variables adjusted correctly in the analysis? | No |
| 23. Has a loss-to-follow impact analysis been done? | Not applicable |
| 24. Were any important primary outcomes missing? | Yes |
| 25. Were the statistical methods adequate? | Yes |
| 26. Were any expected adverse events missing from the results? | Yes |
| 27. Was the statistical analysis of adverse events adequate? | No |
| 28. Are the results reliable considering the limitations of the study? | Yes |
| 29. Has the funding source been identified? | No |
| **Risk of general bias** | **High** |
